# Malaria Risk among Internally Mobile Individuals and Heterogeneous Mobility Patterns in Two Hypoendemic Communities: Implications for Malaria Elimination in the Peruvian Amazon

**DOI:** 10.64898/2026.06.10.26355294

**Authors:** Roberson Ramirez, Carlos Acosta, Pamela Rodríguez, Luis Cabrera-Sosa, Ananias A. Escalante, Joseph M. Vinetz, Katherine Torres, Dionicia Gamboa

## Abstract

**Background:** Human mobility is increasingly recognized as a key factor influencing malaria transmission dynamics, particularly in low-transmission settings approaching elimination. This study aimed to assess mobility patterns and their association with malaria risk in two hypoendemic communities in the Peruvian Amazon.

**Method:** A longitudinal study was conducted in the communities of Libertad and Urcomiraño (Mazán River basin). Monthly population screenings were combined with weekly active and passive case detection. A total of 678 individuals were enrolled. Mobility patterns were assessed through structured questionnaires, and social network analysis was used to characterize travel connections. Log-binomial regression analysis was applied to identify risk factors associated with malaria infection.

**Result:** Internally, mobile individuals in Libertad showed a higher malaria incidence (>32.47 cases per 1,000 person-months) than those in Urcomiraño (<10.15 cases per 1,000 person-months). Travel networks were mainly connected to Mazan district and Iquitos city, followed by local streams such as Armas and Arahuana. Mobility was primarily driven by family, administrative and occupational activities. Male sex (PR = 2.15, 95% CI: 1.37 - 3.37) and age ≥15 years (PR = 1.98, 95% CI: 1.24 - 3.19) were significantly associated with malaria infection (p-value < 0.05).

**Conclusion:** Internally mobile populations represent a key high-risk group sustaining malaria transmission in hypoendemic settings. Targeted interventions focusing on mobile individuals should be integrated into malaria elimination strategies in the Peruvian Amazon and similar endemic regions.

## Introduction

Malaria is a major infectious disease prevalent in tropical and subtropical countries. According to the WHO, during 2023, 263 million malaria cases were reported worldwide, and in the Americas, malaria cases have declined by approximately 65.4% between 2015 and 2022 (1). In Peru, more than 90% of malaria cases are diagnosed in the Loreto Region. In 2021, the Ministry of Health from Perú reported nearly 19,000 cases. By 2023, cases had increased up to 33,000, with *P. vivax* being more prevalent (82%) (2).

Malaria transmission in the Peruvian Amazon is linked to the presence of competent *Anopheles* vectors, especially *Nyssorhynchus darlingi* (3,4). Additionally, infections caused by *Plasmodium* spp. are characterized mostly as submicroscopic and asymptomatic in communities close to Iquitos (5,6), with high parasitemia in remote areas (i.e., Nueva Jerusalen (7)) and low parasitemia in others, such as Santa Emilia (8). These kinds of infections pose a challenge to malaria control plans in the Amazon rainforest.

Malaria transmission is heterogeneous. Whereas a vector’s flight range is typically less than 500 meters, multiple factors drive changes in exposure. For instance, studies have shown that the changes in the environment, such as deforestation, increase exposure of humans to vectors (3,9), while social-economic factors such as maternal employment status and education level, household infrastructure (construction material of walls, roof, and floor of house, wall openings, etc.), household overcrowding, not using bed nets, sharing living spaces with livestock, the distance between the mosquito-breeding site and living house also play a significant role (6,10–13). Therefore, these factors make transmission resilient to interventions, hindering the prospects for eliminating malaria in the future.

Additionally, human mobility, particularly occupation-related travel, has been identified as a factor in the maintenance and expansion of malaria (5,14–16). Recent studies conducted in Peru and in northern and southern Zambia, employing GPS-based approaches, have identified areas of intensive human movement characterized by occupation-related high-connectivity nodes. These regions exhibited diverse mobility patterns, including frequent travel into and out of areas targeted for indoor residual spraying (IRS) and seasonal movement dynamics, with a marked increase in long-distance travel during the dry season (17–19). Other approaches based on mobile phone data showed heterogeneous human movement patterns in Ethiopia, with high mobility during mosquito biting hours in the malaria transmission season due to cross-border movements, internal economic migration linked to agricultural and mining activities, and seasonal population influx into malaria endemic regions (20), while in Kenya enabled the construction of mobility networks and the development of malaria risk maps, suggesting that human movement drives transmission in areas of low endemicity as expected (21).

Finally, epidemiological studies conducted in Bangladesh, Saudi Arabia, Brazil, and Peru have contributed to characterizing spatial and temporal patterns of human mobility, identifying heterogeneous movement patterns and risk groups among internally mobile individuals across short- and long-distance ranges toward high transmission areas (14,22–24). However, further refinements in epidemiological study design and survey instruments tailored to internally mobile populations would strengthen the evidence base in this field. Here, we evaluate a longitudinal epidemiological design to document the impact of mobility on malaria risk in an area of the Peruvian Amazon.

The Plan for the Elimination of Malaria in Peru, launched by the Ministry of Health (MINSA) in 2022, aims to reduce malaria cases by nearly 90% by 2030 through a community-based approach. The high level of human mobility between river basins or communities creates areas with a highly dispersed populations (25), making an ideal setting for this study. The Mazan district, located in the Loreto region, is a hotspot for malaria transmission due to its connections to an extensive river network and high levels of population movement associated with fishing, logging, and agriculture (17,22). Additionally, incidence increased significantly by 88% from 2021 to 2022 (26). Previous studies in the area have documented that human mobility in sustaining malaria transmission, highlighting that travel for work often exposes individuals to higher transmission zones (5,17,22). Still, more granular information is needed to tailor interventions locally. To develop methodologies that can yield the required data, a longitudinal epidemiological design with time-varying mobility exposure was implemented in the Libertad and Urcomiraño communities in Mazan.

## Methods

### Ethics statement

Individuals ≥ 3 months were enrolled after written informed assent/consent was signed by the individual, a parent, or a tutor. For the sample collection, the protocol (“Transmission Dynamics of Residual and Re-emerging Malaria in Amazonia: Defining Pathways Towards Malaria Elimination”) and the assent/consent were reviewed and approved by the Ethics Committee from Universidad Peruana Cayetano Heredia (SIDISI 101518). The data analysis in this study was also approved by that Ethics Committee (SIDISI 207483).

### Study area and population

The study was conducted in the Mazan district, located in Maynas province in Loreto region. Mazan has a surface area of 10082.00 km². Within the Mazan district, the communities of Libertad (3.496° S, 73.234° W) and Urcomiraño (3.361° S, 73.064° W) are located (Figure 1). Libertad and Urcomiraño were selected because of their similar epidemiological characteristics and population size according to previous studies (5,22), but located in different river basins (Mazan and Napo, respectively).

**Figure 1.**
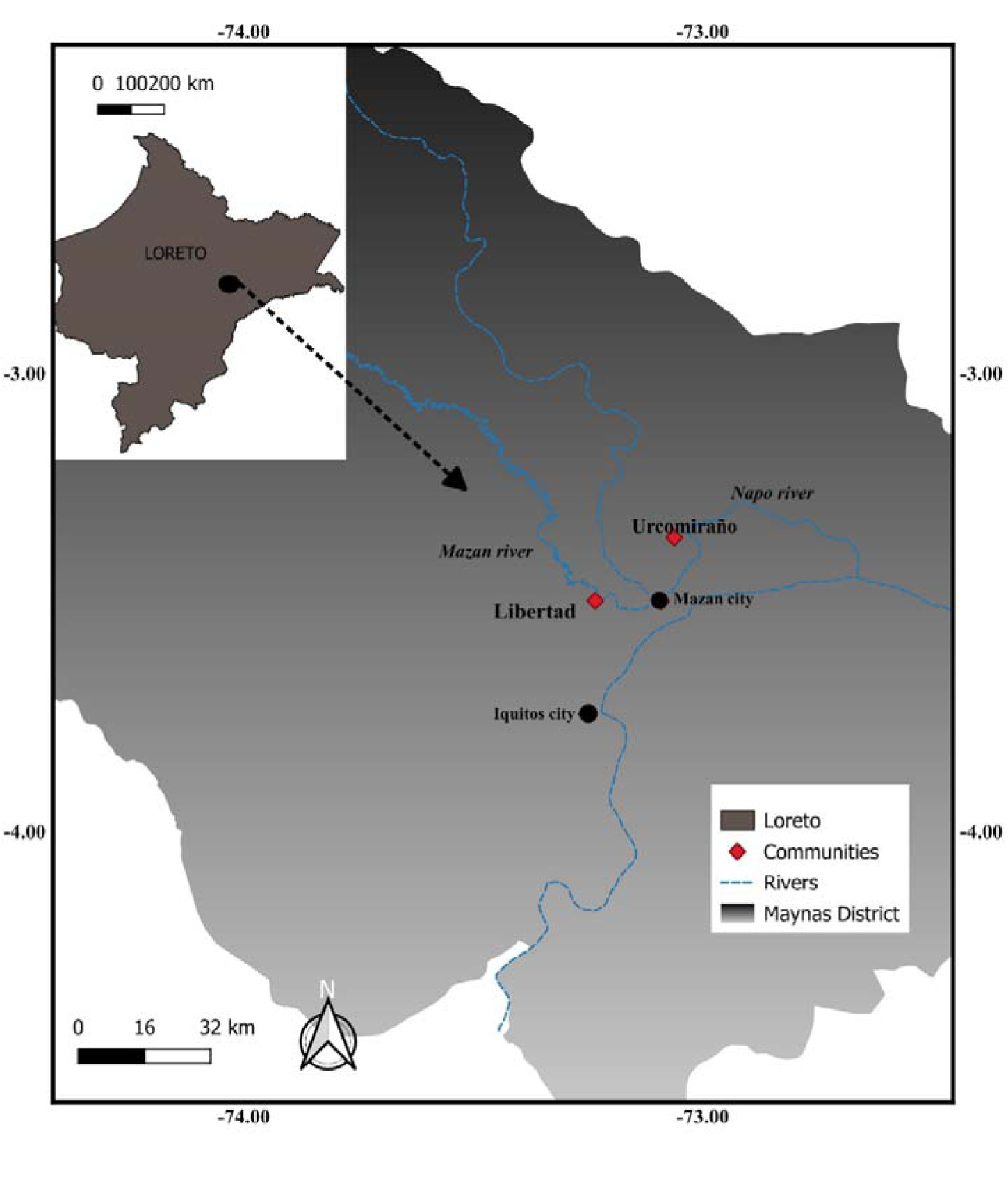
Map of the study area. The map shows the geographical location of the Libertad and Urcomiraño communities, located in the province of Maynas, Loreto Region, Peru. The map was produced using QGIS software version 3.40.7. The cartographic layers in .shp format corresponding to administrative boundaries (Region, province and district) were obtained from the National Institute of Statistics and informatics (INEI), while the hydrological data (river network) were downloaded from the National Water Authority (ANA).

The only access to both communities is by river. Libertad and Urcomiraño lack access to electricity and drinking water; their main economic activities are agriculture, fishing, commerce, and wood extraction. Additionally, each community has a health center serving its population. The weather in the Mazan district is tropical, with an annual temperature range of 31.1 – 21.95 °C, relative humidity of 80%, and an average annual rainfall ranging between 2,800 and 3,200 mm, with periods of flooding during the months of November to June and unflooding in August to October (27).

In this study, an internally mobile individual was defined as a resident who had spent at least one night outside his or her home community within the same geographic region during the preceding month, regardless of the purpose of displacement, and a symptomatic individual was one who presented at least one among fever, headache, or chills.

Data and capillary blood samples were collected and assessed from the entire study community, with a coverage of 98%. In Libertad, data were collected from 56 houses with a population of 358 individuals, and in Urcomiraño, from 59 houses with a population of 320 individuals.

On the other hand, during the active case detection week, a total of 144 internally mobile individuals were identified across the two study communities. Information on these individuals was used to describe the social network analysis, frequency, and purpose of displacement.

### Study design and data collection

Samples and population data were collected over two consecutive years (February–August 2021 and February–August 2022) in the Libertad and Urcomiraño communities. Socio-demographic information was collected through epidemiological surveys. Each participant provided between 1 and 12 samples or records. At the same time, active case detection was carried out on a weekly basis to search each household for individuals with or without malaria symptoms who had traveled in the last month (Figure 2). Internal mobility was assessed by asking household members whether any resident had spent at least one night away from the community during the preceding month, or whether any visitor had arrived at the household within the same period.

**Figure 2.**
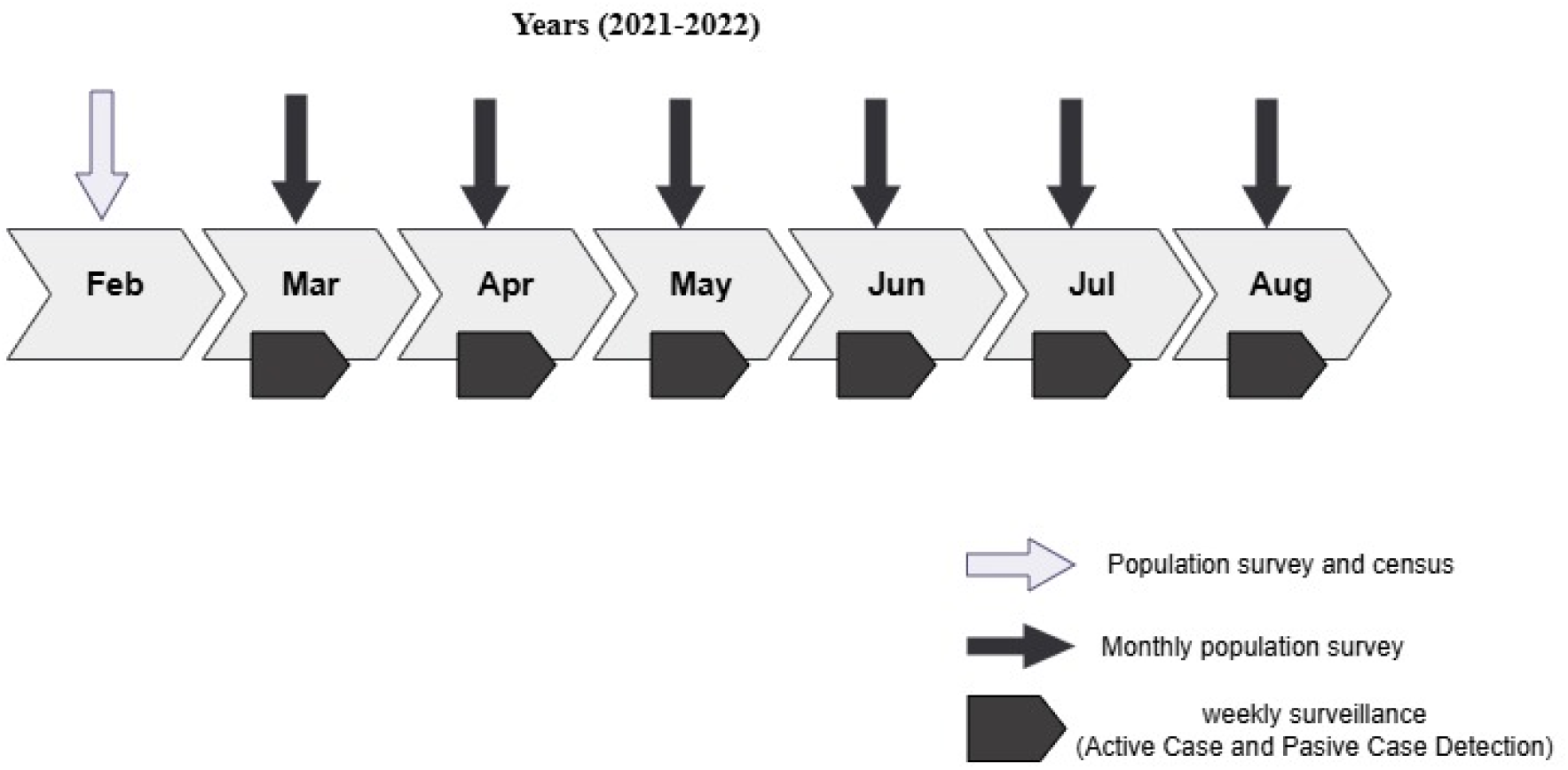
Study and sample collection design. The samples and data were collected from February and August in 2021 and 2022. Active case detection and passive case detection were the strategies used during weekly surveillance. Finally, monthly population surveys in Libertad and Urcomiraño were also conducted.

Finally, passive case detection was used in the health centers of each community to identify symptomatic individuals who could not be found at home during the population/community surveys. Capillary blood was collected in EDTA microtainer tubes (Vacutest, Kima ^®^, Padua, Italy) y and stored at -20 °C until processing in the laboratory.

### Microscopic and molecular diagnosis of malaria

Microscopic diagnosis was performed in accordance with MINSA guidelines (28). Microscopic diagnosis was performed monthly during the survey and in active and passive surveillance of cases. Individuals with microscopic positive results for malaria were reported to the health center, where they received treatment according to MINSA guidelines.

Molecular diagnosis was performed retrospectively. Parasite DNA was extracted from blood cells using EZNA^®^ Blood DNA kit (Omega BIO-TEK, GA, USA). DNA from each sample was resuspended in 50μL of elution buffer. The qPCR protocol proposed by Rougemont et al. 2004 (29) was used to differentiate *Plasmodium* species, employing specific probes for *P. vivax* and *P. falciparum* (29). Parasitemia was determined based on a standard curve, with five serial dilutions in 1/10 factor (2000 to 0.2 p/μL). The detection limit of the protocol was 2 parasites/μL (29).

### Description of population and human mobilization

Information from each participant was recorded through a sociodemographic survey using the digital tool Open Data Kit (ODK) on tablet devices. Upon completion, the surveys were uploaded to a digital database (REDCap). The recorded data was downloaded in Excel format and analyzed.

For cleaning and handling the epidemiological database from the monthly sweeps and the internally mobile individuals survey, the statistical program R version 4.2.3 and RStudio were used. The *tidyverse*, *dplyr*, and *sjmisc* packages were employed. The epidemiological description of participants and internally mobile individuals from both communities was carried out in R using the *stats*, *readxls*, *tidyverse*, *writexl*, and *dplyr* packages. For statistical significance, a p-value < 0.05 was used. Fisher’s exact tests and Chi-square tests were applied to compare the proportions of variables between communities

### Prevalence and Incidence rates by community, year, and travel exposure status analysis

Annual prevalence was estimated as the proportion of individuals with at least one PCR-confirmed positive result within each calendar year and community. Individuals were classified as prevalent cases if they had at least one positive PCR result during the year. The denominator comprised all individuals with available PCR data for the corresponding year. Ninety-five percent confidence intervals (95% CIs) were calculated using exact binomial methods.

Incidence rates among internally mobile and non-internally mobile individuals were estimated using a person-time approach, with person-months as the unit of analysis. Each individual contributed one person-month at risk per calendar month under observation. If an individual tested positive more than once within the same calendar month, only the first positive result was counted as an incident case.

Internally mobile individual exposure was treated as a time-varying variable defined at the monthly level. Incident cases were defined as PCR-confirmed positive results occurring within a given month, and incidence rates were calculated as the number of incident cases per 1,000 person-months. In this case, 95% CIs were estimated assuming a Poisson distribution.

### Malaria risk factors analysis

Log-binomial regression analysis with Firth correction was conducted to identify factors associated with malaria infection, in order to reduce small-sample bias and address potential convergence issues due to sparse data. Prevalence ratios (PRs) and their 95% CIs were estimated for each variable of interest. Initially, initial models (PR¹) were constructed to evaluate the individual association of each factor with infection. Subsequently, an adjusted model (PR²) was developed using a backward selection procedure based on the Akaike Information Criterion (AIC) to retain the most relevant variables.

Variables included in the analysis were occupation, recent internal mobility in the preceding month, sex, and age group (<15 years or ≥15 years). A p-value < 0.05 was considered statistically significant.

### Social network analysis

A social network analysis was performed to characterize travel connections among the surveyed communities. Each node represented a community, and each edge indicated reported travel between two communities. Edge weights corresponded to the frequency of reported trips, and node size was proportional to the degree (number of direct connections). The network was constructed and analyzed using the *igraph* package in R (version 4.5.1). Descriptive network metrics, including degree, edge weight, and network density, were calculated to identify highly connected communities (hubs). Geographic coordinates of each community were used to map the spatial distribution of connections using *ggplot2* and *sf* packages. Networks were categorized by travel frequency: low (1-6), medium (7-12), high (13-18) to visualize the relative intensity of mobility patterns across the study area. The categorization was performed using the equal-interval classification method, in which the total range of variable values is divided into a fixed number of equal-sized intervals.

Data cleaning, processing, and all statistical and network analyses were conducted using R (version 4.5.1). The *igraph* package was used for network analysis. A two-sided p-value < 0.05 was considered statistically significant.

## Results

### Sociodemographic characteristics of the study participants

Sociodemographic data were collected from 678 individuals, distributed between Libertad (n = 358) and Urcomiraño (n = 320) communities. Individuals were predominantly male (54.3%) and adults (> 15 years, 59.9%), with no significant differences between communities for either variable. Education level differed significantly between communities (p = 0.001), with Urcomiraño showing a higher proportion of individuals with secondary or higher education (36.3%) compared to Libertad (23.7%). Regarding time of residence, significant differences were also observed (p = 0.001), with Urcomiraño having a higher proportion of long-term residents (≥10 years: 51.2%) than Libertad (8.7%). Malaria history likewise differed between communities (p = 0.007): Libertad had a higher proportion of individuals with more than four lifetime episodes (34.6% vs. 21.9%). No significant differences were found between communities in terms of occupation, bed net use, or household characteristics (Table 1).

**Table 1:** Population and Household Characteristics of Libertad and Urcomiraño communities.

|  | Libertad |  | Urcomiraño |  | Total |  |  |
| --- | --- | --- | --- | --- | --- | --- | --- |
|  | n = 358 | % | n = 320 | % | 678 | % | p-value |
| **Gender** |  |  |  |  |  |  |  |
| Male | 192 | 53.6 | 176 | 55.0 | 368 | 54.3 | 0.780 |
| Female | 166 | 46.4 | 144 | 45.0 | 310 | 45.7 |  |
| **Age** |  |  |  |  |  |  |  |
| ≥15 | 215 | 60.1 | 191 | 59.7 | 406 | 59.9 | 0.984 |
| <15 | 143 | 39.9 | 129 | 40.3 | 272 | 40.1 |  |
| **Education level (≥ 3 y)\*** |  |  |  |  |  |  |  |
| None | 70 | 19.6 | 62 | 19.4 | 132 | 19.5 | 0.001 |
| Initial or primary school | 203 | 56.7 | 142 | 44.4 | 345 | 50.9 |  |
| Secondary or high | 85 | 23.7 | 116 | 36.3 | 201 | 29.6 |  |
| **Main occupation**<br>(Logger, fisher or farmer) |  |  |  |  |  |  |  |
| No | 203 | 56.7 | 191 | 59.7 | 394 | 58.1 | 0.479 |
| Yes | 155 | 43.3 | 129 | 40.3 | 284 | 41.9 |  |
| **Time in the village (age > 10)\*** |  |  |  |  |  |  |  |
|  | n= 117 |  | n=41 |  | n= 158 |  |  |
| < 2 | 54 | 15.1 | 8 | 19.5 | 62 | 39.2 | 0.001 |
| 2 - 9.9 | 32 | 8.9 | 12 | 29.3 | 44 | 27.8 |  |
| ≥ 10 | 31 | 8.7 | 21 | 51.2 | 52 | 32.9 |  |
| **Malaria episodes throughout life\*** |  |  |  |  |  |  |  |
| 0 | 81 | 22.6 | 83 | 25.9 | 164 | 24.2 | 0.007 |
| 1 | 56 | 15.6 | 65 | 20.3 | 121 | 17.8 |  |
| 2 to 3 | 97 | 27.1 | 102 | 31.9 | 199 | 29.4 |  |
| > 4 | 124 | 34.6 | 70 | 21.9 | 194 | 28.6 |  |
| **Use net in the last three night** |  |  |  |  |  |  |  |
| Yes | 355 | 99.2 | 314 | 98.1 | 669 | 98.7 | 0.319 |
| No | 3 | 0.8 | 6 | 1.9 | 9 | 1.3 |  |
| **Household Characteristics** |  |  |  |  |  |  |  |
| **Wall material** |  |  |  |  |  |  |  |
| Wood | 351 | 98.0 | 311 | 97.2 | 662 | 97.6 | 0.614 |
| Brick, cement and other | 7 | 2.0 | 9 | 2.8 | 16 | 2.4 |  |
| **Floor material** |  |  |  |  |  |  |  |
| Wood | 322 | 89.9 | 298 | 93.1 | 620 | 91.4 | 0.169 |
| Dirt | 36 | 10.1 | 22 | 6.9 | 58 | 8.6 |  |

\*Significance difference, y = years.

### Prevalence, Incidence rates by community, year, and travel exposure status

During 2021 and 2022, the prevalence by qPCR in Libertad was 0.06 (95% CI: 0.04 - 0.09) and 0.10 (95%: CI 0.07 - 0.14), respectively. However, Urcomiraño presented a low prevalence during the same period of the study: 0.03 (95% CI: 0.01 - 0.05) and 0.01 (95% CI: 0 - 0.04), respectively.

Figure 3 illustrates that in Libertad community, internally mobile individuals experienced approximately twice the risk of malaria compared with non-internally mobile individuals during 2021. This pattern was reflected in a markedly higher incidence rate among internally mobile individuals than among their non-mobile counterparts (38.46 vs. 19.03 cases per 1,000 person-months), supporting the interpretation that imported infections played a substantial role in malaria transmission (p-value = 0.064). In 2022, however, incidence among non-internally mobile individuals increased considerably (35.53 cases per 1,000 person-months), reaching comparable levels to those among internally mobile individuals (32.47 cases per 1,000 person-months) (p-value = 0.999).

**Figure 3.**
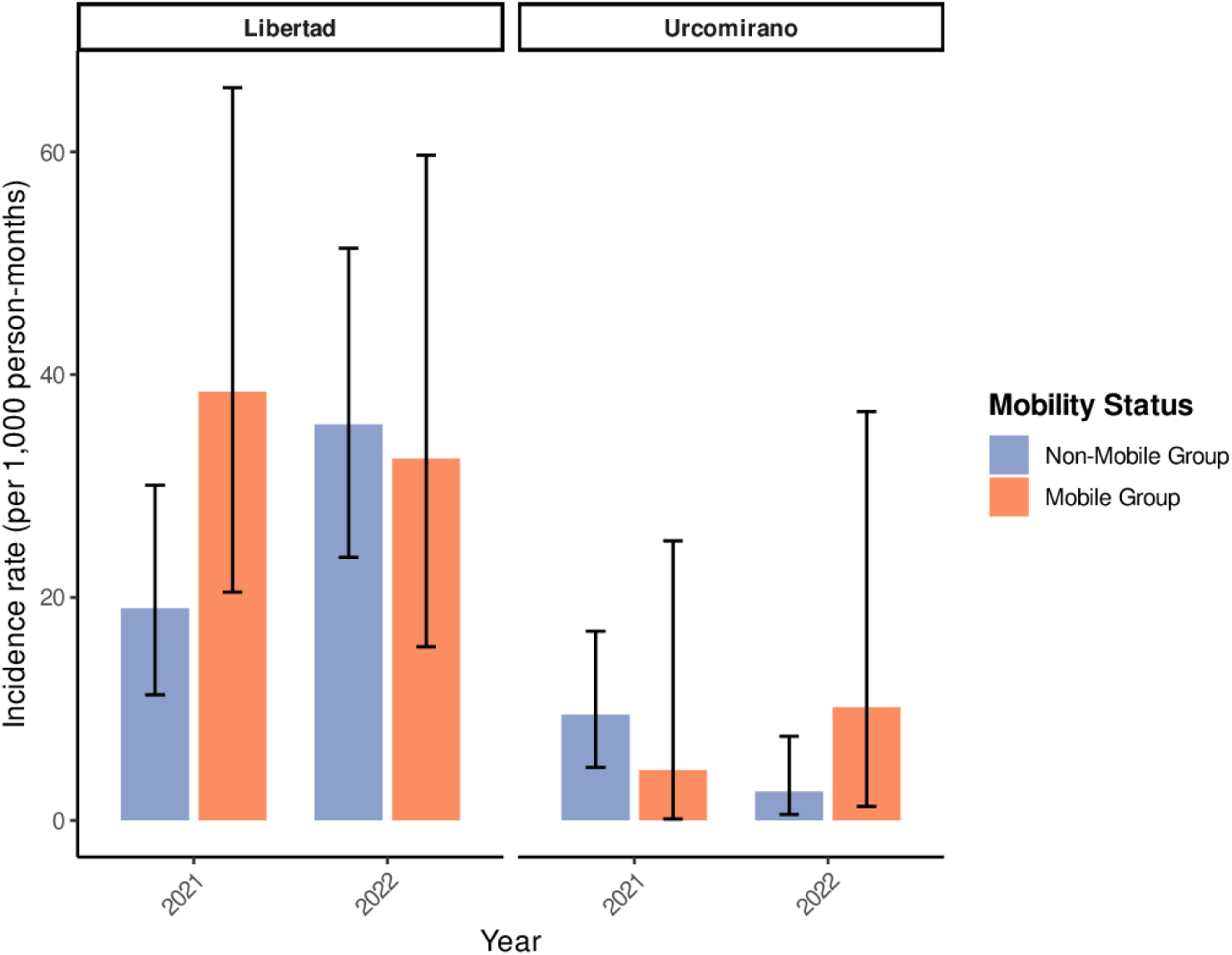
Incidence rates in Libertad and Urcomiraño by year and mobility status. Incidence rates of PCR-confirmed infection per 1,000 person-months among mobile and no-mobile group exposure across calendar years, stratified by community. Bars represent incidence rate estimates, and error bars indicate 95% CIs.

In Urcomiraño, a different temporal pattern was observed in 2021. Non-internally mobile individuals showed higher incidence rates than internally mobile individuals (9.48 vs. 4.50 cases per 1,000 person-months), indicating predominantly local transmission (p-value = 0.7044). By contrast, in 2022, incidence rates increased among internally mobile individuals than among their non-mobile counterparts (10.15 vs. 2.58 cases per 1,000 person-months (p-value = 0.1556), suggesting that human mobility may have contributed to the persistence of malaria transmission in this community.

Across both study years, Libertad consistently exhibited higher malaria incidence rates among internally mobile individuals than Urcomiraño, highlighting a sustained vulnerability of mobile populations and underscoring the role of human movement in shaping heterogeneous malaria transmission dynamics in these Amazonian communities (Supplementary Table 1).

### Mobility patterns of internally mobile individuals from the communities of Libertad and Urcomiraño

The social network analysis revealed a heterogeneous structure of travel connections among the surveyed communities (Figure 4). Urcomiraño and Libertad emerged as the main hubs, exhibiting the highest degree of connectivity and travel frequency. The overall network (Figure 4A) showed dense interconnections centered around these nodes, with varying link intensities representing different travel frequencies. The geographic representation of the main connections (Figure 4B) highlighted the central position of Iquitos as a strong linkage among nearby communities. Connections associated with Urcomiraño (Figure 4C) extended predominantly toward southern and peripheral areas as Chasute and Sucusari, whereas those linked to Libertad (Figure 4D) showed a more clustered pattern involving nearby settlements as Antioquia, Muena Grande, Santa Cruz, Yutococha, San Jose de Yanashi, and Maniti. These findings suggest that human mobility in the study area is spatially structured around a few highly connected communities that may play a key role in maintaining regional connectivity. A median (range: 7–12) travel frequency was also observed from Urcomiraño to the Armas stream, and similarly from Libertad to the Arahuana stream.

**Figure 4.**
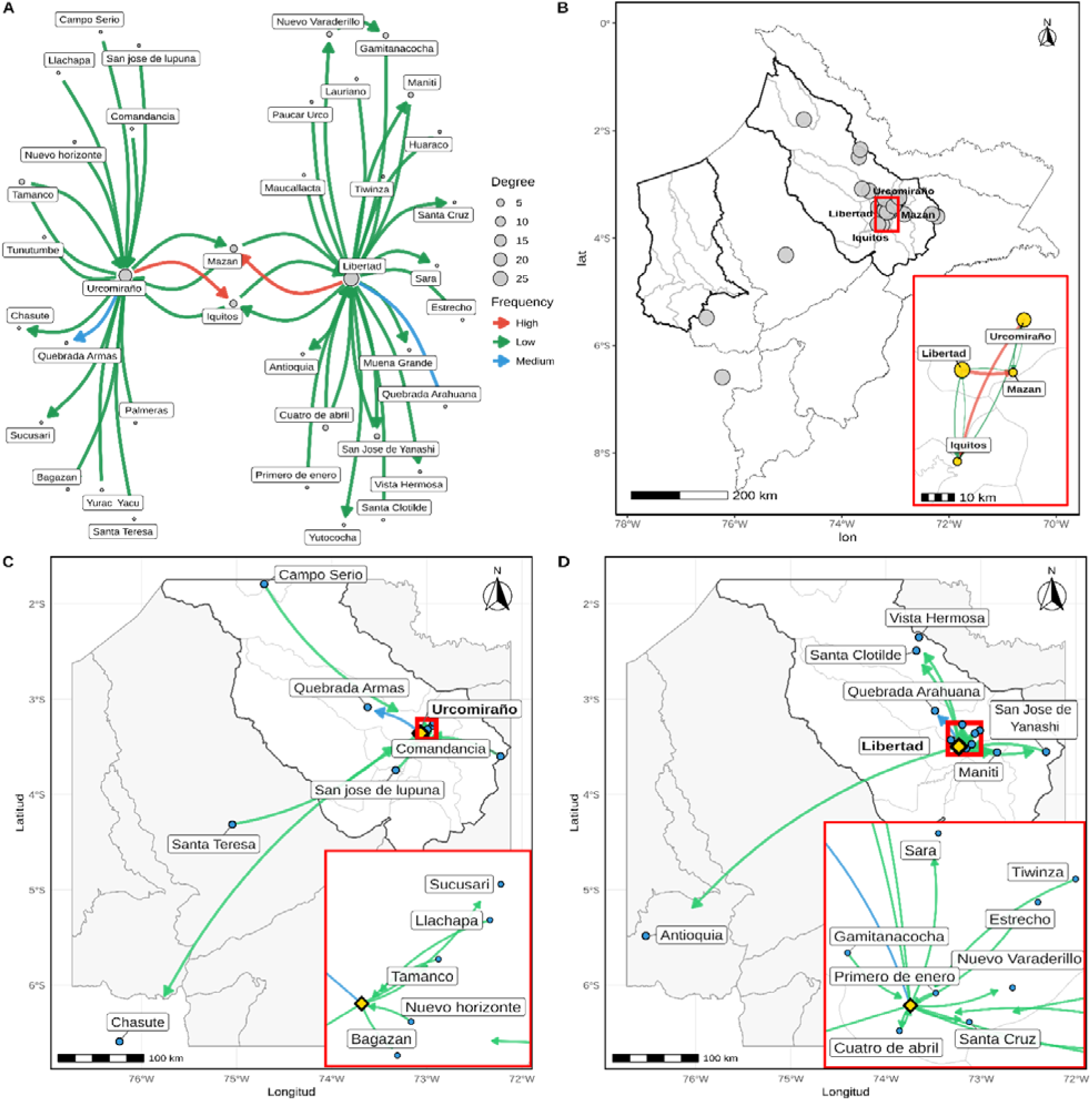
Social network analysis of Internally Mobile Individuals connections among study sites. (A) Network diagram showing the degree and travel frequency categorized into three levels (high, medium, and low). (B) Geographic location of the main Internally Mobile Individuals connections. (C) Geographic distribution of the connections associated with **Urcomiraño**. (D) Geographic distribution of the connections associated with **Libertad**

On the other hand, the primary reasons for displacement were family matters (44%) and work/trade in both communities. In Ercolino, purchasing food represented an additional reason for mobility (Supplementary Figure 1).

### Risk factors associated with malaria

In the initial analysis (Table 2), working in agriculture, forestry, or fishing was significantly associated with higher malaria prevalence compared to other occupations (PR = 1.97; 95% CI:1.13 – 3.44; p-value = 0.017). Recent internal mobility within the preceding month was also associated with increased risk of infection (PR = 1.76; 95% CI: 1.12 – 2.77; p-value = 0.014). Males had a higher prevalence of malaria than females (PR= 2.13; 95% CI: 1.35 – 3.35; p-value = 0.001). Additionally, participants aged 15 years or older had a higher risk than those under 15 years (PR = 2.07; 95% CI: 1.29 – 3.31; p-value = 0.002).

**Table 2.**
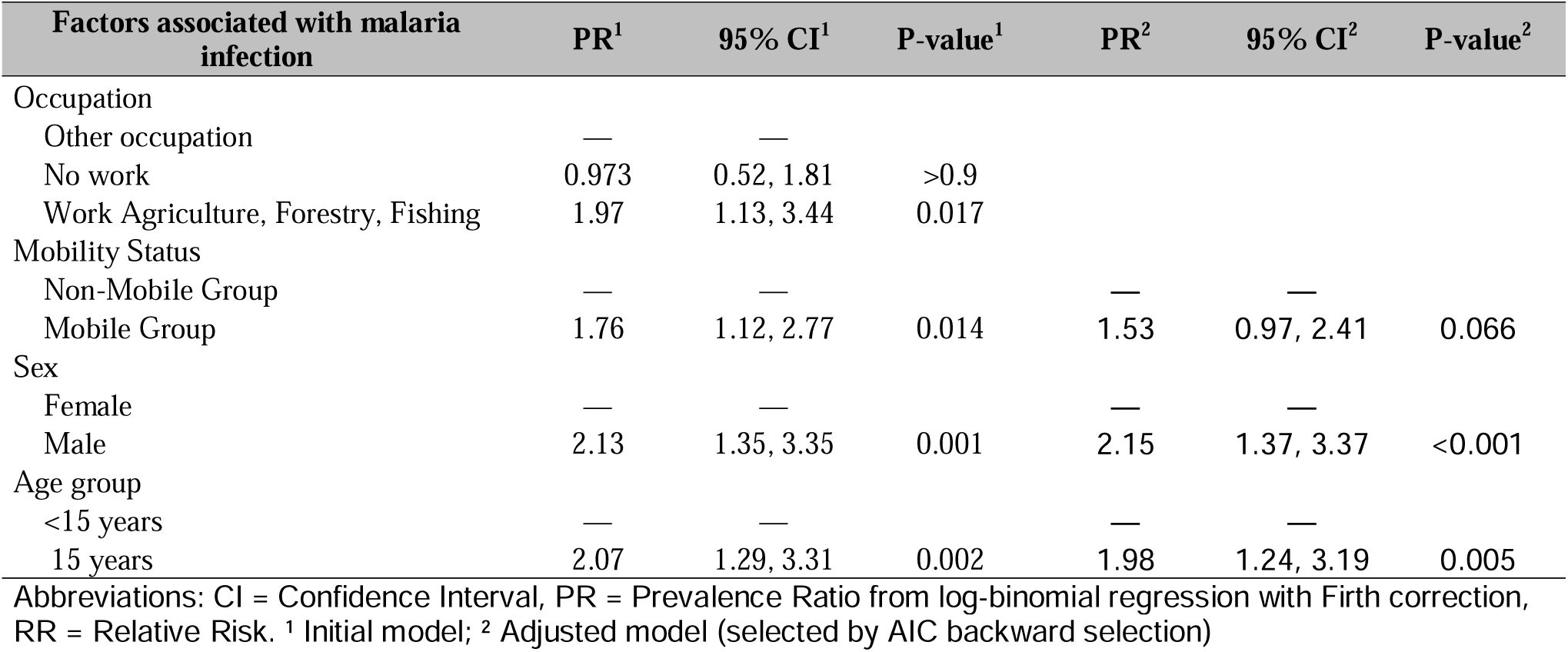
Factors associated with malaria infection. Initial (PR¹) and adjusted (PR²) prevalence ratios with 95% confidence intervals (CI) and corresponding p-values are presented. The initial model includes univariate associations of each factor with malaria infection, while the adjusted model was selected using backward stepwise selection based on the Akaike Information Criterion (AIC). Occupation, recent travel in the last month, sex, and age group were analyzed as potential predictors. Abbreviations: CI = Confidence Interval; PR = Prevalence Ratio from log-binomial regression with Firth correction; RR = Relative Risk

| Factors associated with malaria infection | PR<sup>1</sup> | 95% CI<sup>1</sup> | P-value<sup>1</sup> | PR<sup>2</sup> | 95% CI<sup>2</sup> | P-value<sup>2</sup> |
| --- | --- | --- | --- | --- | --- | --- |
| Occupation |  |  |  |  |  |  |
| Other occupation | — | — |  |  |  |  |
| No work | 0.973 | 0.52, 1.81 | >0.9 |  |  |  |
| Work Agriculture, Forestry, Fishing | 1.97 | 1.13, 3.44 | 0.017 |  |  |  |
| Mobility Status |  |  |  |  |  |  |
| Non-Mobile Group | — | — |  | — | — |  |
| Mobile Group | 1.76 | 1.12, 2.77 | 0.014 | 1.53 | 0.97, 2.41 | 0.066 |
| Sex |  |  |  |  |  |  |
| Female | — | — |  | — | — |  |
| Male | 2.13 | 1.35, 3.35 | 0.001 | 2.15 | 1.37, 3.37 | <0.001 |
| Age group |  |  |  |  |  |  |
| <15 years | — | — |  | — | — |  |
| 15 years | 2.07 | 1.29, 3.31 | 0.002 | 1.98 | 1.24, 3.19 | 0.005 |

Abbreviations: CI = Confidence Interval, PR = Prevalence Ratio from log-binomial regression with Firth correction, RR = Relative Risk. <sup>1</sup> Initial model; <sup>2</sup> Adjusted model (selected by AIC backward selection)

In the adjusted model, which included variables selected based on AIC, the associations remained in the same direction. Working in agriculture, forestry, or fishing showed an effect on infection, without reaching a statistical significance in the final model. Recent internal mobility tended to increase malaria prevalence, but the effect was marginally significant (PR = 1.53; 95% CI: 0.97 - 2.41; p-value = 0.066). Male sex remained significantly associated with a higher prevalence of infection (PR = 2.15; 95% CI:1.37 - 3.37; p-value < 0.001), as did the age group ≥15 years (PR = 1.98; 95% CI: 1.24 - 3.19; p-value = 0.005).

## Discussion

In this study, the mobility pattern observed in Libertad and Urcomiraño communities was characterized by a heterogeneous structure of travel connections. We observed a high to moderate frequency of displacement from Libertad to the town of Mazan, where most of the economic activity of the Mazan River basin is concentrated, and to the Arahuana stream. In contrast, in Urcomiraño, most internally mobile individuals traveled to the city of Iquitos (the Capital of Loreto) and to the Armas stream.

Similar patterns have been reported in other communities of the Mazan district (17,22,30), where nearby communities, densely populated centers (the towns of Mazan and Indiana), and rural campsites were identified as the main travel destinations. Travel among internally mobile individuals was mainly driven by family-related reasons, administrative procedures, and work or trade (Supplementary Figure 1).

Internally mobile individuals from Libertad consistently exhibited higher malaria risk (>32 cases per 1,000 person-months) than those from Urcomiraño, with incidence rate ratios suggesting a stable or increasing trend in 2022 compared to 2021, these patterns of transmission may reflect the gradual restoration of human mobility from COVID-19-related restrictions, which may have contributed to the increase in malaria transmission observed in other communities throughout the Peruvian Amazon (31). Internally mobile individuals remained an important group in maintaining malaria transmission compared with non-internally mobile individuals. Similarly, internal local movements within this group may help explain locally acquired cases. These findings also confirm that short-distance travel to higher-transmission areas is epidemiologically relevant, comparable in importance to international travel (32).

These findings support the hypothesis that human mobility contributes to the persistence of malaria transmission in rural communities of the Peruvian Amazon through imported infections and connectivity between endemic areas. This interpretation aligns with recent evidence that human mobility creates a high connection between rural and peri-urban areas and supports the movement of parasites across the landscape of the Amazon Rainforest in Peru, linking the complex transmission patterns (5,19,22).

In Urcomiraño, travel patterns were characterized by longer distance movements, whereas in Libertad, mobility was primarily local. This suggests that the communities settled along the Mazan and Napo river basins may function as a single transmission focus, likely driven by long-standing socio-geographical connections. Supporting this, similar parasite lineages have been reported across these communities (33); however, population genetic studies are required to formally confirm this assumption.

We considered that heterogeneous patterns of human mobility shape the heterogeneity of malaria transmission, providing a plausible explanation for the local micro-epidemiological characteristics observed in endemic settings (5). These findings reinforce the growing body of evidence suggesting that fine-scale human movement should be explicitly considered when designing and optimizing malaria control and elimination strategies, particularly in highly connected rural and riverine environments(5,6).

Additionally, previous studies have suggested that the connectivity generated by internally mobile individuals can create biological corridors (33–35), facilitating parasite movement from a source to a sink. For instance, a moderate frequency of travel to the Arahuana and Armas streams for economic and recreational activities likely increased their exposure to mosquito-borne infections. Although these locations received 7.9% and 5.8% of all incoming travel, they were simply destinations people visited, but did not continue on to other places from there. As a result, they played a minor role in connecting the region. On the other hand, Libertad and Urcomiraño were not only frequently visited but also acted as travel hubs, where people passed through on their way to other communities (scores of 0.149 and 0.146, respectively), making them key points of regional connection) (Supplementary Table 2).

We confirm that internally mobile individuals represent a previously reported risk factor for malaria transmission (22,36,37), and therefore recommend that hybrid strategies should be implemented. For instance, combining interventions targeting local transmission (indoor residual spraying, insecticide-treated nets, and other vector control measures), while also focusing on this population for systematic monitoring before and after returning from higher-transmission areas, through the use of rapid diagnostic tests and timely treatment protocols. This study identified adults aged 15 years and older as a higher-risk group, likely due to greater participation in outdoor occupational activities such as farming, fishing, and forestry, which increase exposure to infected mosquitoes. Consistent with previous findings (22,38,39), young men who are engaged in outdoor occupational and social activities commonly have a higher vulnerability to contracting malaria than other populations, indicating that they may have been missed as a source of continuous transmission.

The findings presented in this work were obtained in the context of transitioning out of the mandatory lockdown resulting from the pandemic that emerged in 2020, and therefore, the resumption of daily activities, which could affect the actual picture of human mobilization. We recommend to conduct additional similar studies because this pattern could change.

## Limitations

Finally, this study has several limitations. First, sampling was conducted only between February and August, corresponding to the flooding season; therefore, potential seasonal variations related to non-flooding were not captured. However, mobility is typically higher during this period due to increased river connectivity, and patterns were consistent across two consecutive years. Second, some epidemiological data were lost due to incomplete follow-up among internally mobile individuals, though these cases were excluded from analysis.

## Conclusions

Internally mobile individuals from the communities of Libertad and Urcomiraño remain an important risk group for malaria. These internally mobile individuals facilitate connectivity between distinct transmission zones and exhibit heterogeneous mobility patterns, largely driven by family-related, administrative, and occupational activities. Therefore, this population should be explicitly considered in current malaria elimination plans in Peru, as targeted interventions aimed at reducing the malaria risk associated with internally mobile individual and it may be critical for interrupting ongoing transmission.

## Authors contribution

RR and DG conceived and designed the study. Field site collections coordinated by RR. DG provided guidance on study design, sample selection for molecular and data analysis for this study. PR and CA conducted molecular assays, support on laboratory and analyzed the data respectively. KT, DG and AAE supervised the work. RR and CA wrote the original draft. JMV, LCS, DG, KT and AAE reviewed, edited and supervised the manuscript. All authors have read and agreed to the published version of the manuscript.

## Data Availability

All data produced in the present study are available upon reasonable request to the authors

## Acknowledgments

We would like to acknowledge all study participants and the local authorities of the Libertad and Urcomiraño communities for their enthusiastic support of this research, as well as the field staff of the Amazonian ICEMR project for their dedication during surveys and follow-up activities.

## Supplementary Information

Supplementary Table 1. Incidence rates by community, year, and travel exposure status.

Supplementary Figure 1. Proportion of travel reasons among individuals from the communities of Libertad and Urcomiraño.

Supplementary Figure 2. Internally Mobile Individuals frequency vs. Centrality (PageRank) and structural metrics.

