## Supplementary material for "Malaria Risk among Internally Mobile Individuals and Heterogeneous Mobility Patterns in Two Hypoendemic Communities: Implications for Malaria Elimination in the Peruvian Amazon": Suplemental material

| Community | Year | Mobility exposure | Cases | Person-months | Incidence rate* | 95% CI |
| --- | --- | --- | --- | --- | --- | --- |
| Libertad | 2021 | No | 18 | 946 | 19.03 | 11.28 - 30.07 |
| Libertad | 2021 | Yes | 13 | 338 | 38.46 | 20.48 - 65.77 |
| Libertad | 2022 | No | 28 | 788 | 35.53 | 23.61 - 51.36 |
| Libertad | 2022 | Yes | 10 | 308 | 32.47 | 15.57 - 59.71 |
| Urco Miraño | 2021 | No | 11 | 1160 | 9.48 | 4.73 - 16.97 |
| Urco Miraño | 2021 | Yes | 1 | 222 | 4.50 | 0.11 - 25.1 |
| Urco Miraño | 2022 | No | 3 | 1162 | 2.58 | 0.53 - 7.54 |
| Urco Miraño | 2022 | Yes | 2 | 197 | 10.15 | 1.23 - 36.67 |
| * Incidence rate per 1,000 person-months. | | | | | | |

**Supplementary Table 1**. Incidence rates by community, year, and mobility exposure status.

A)

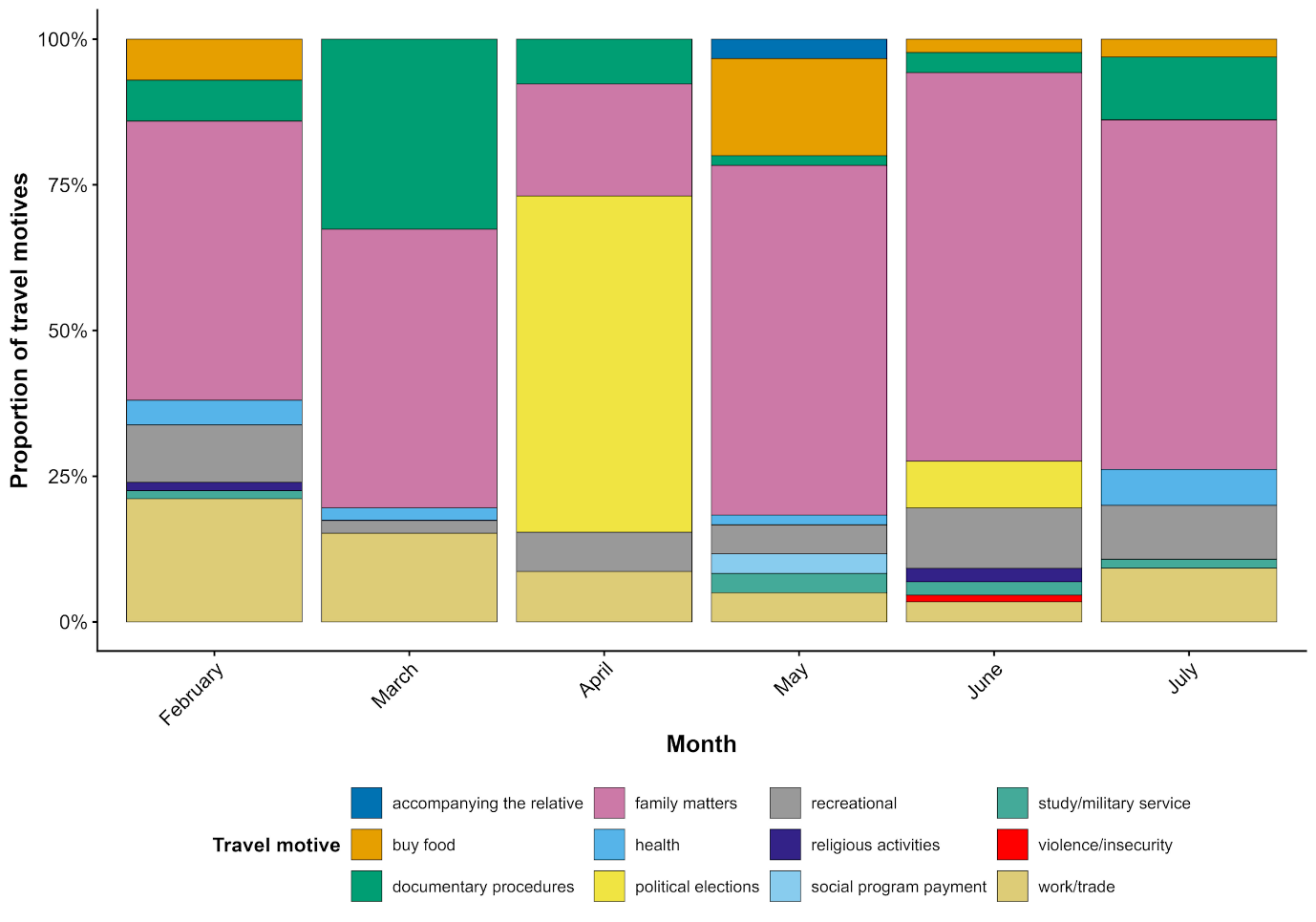

B)

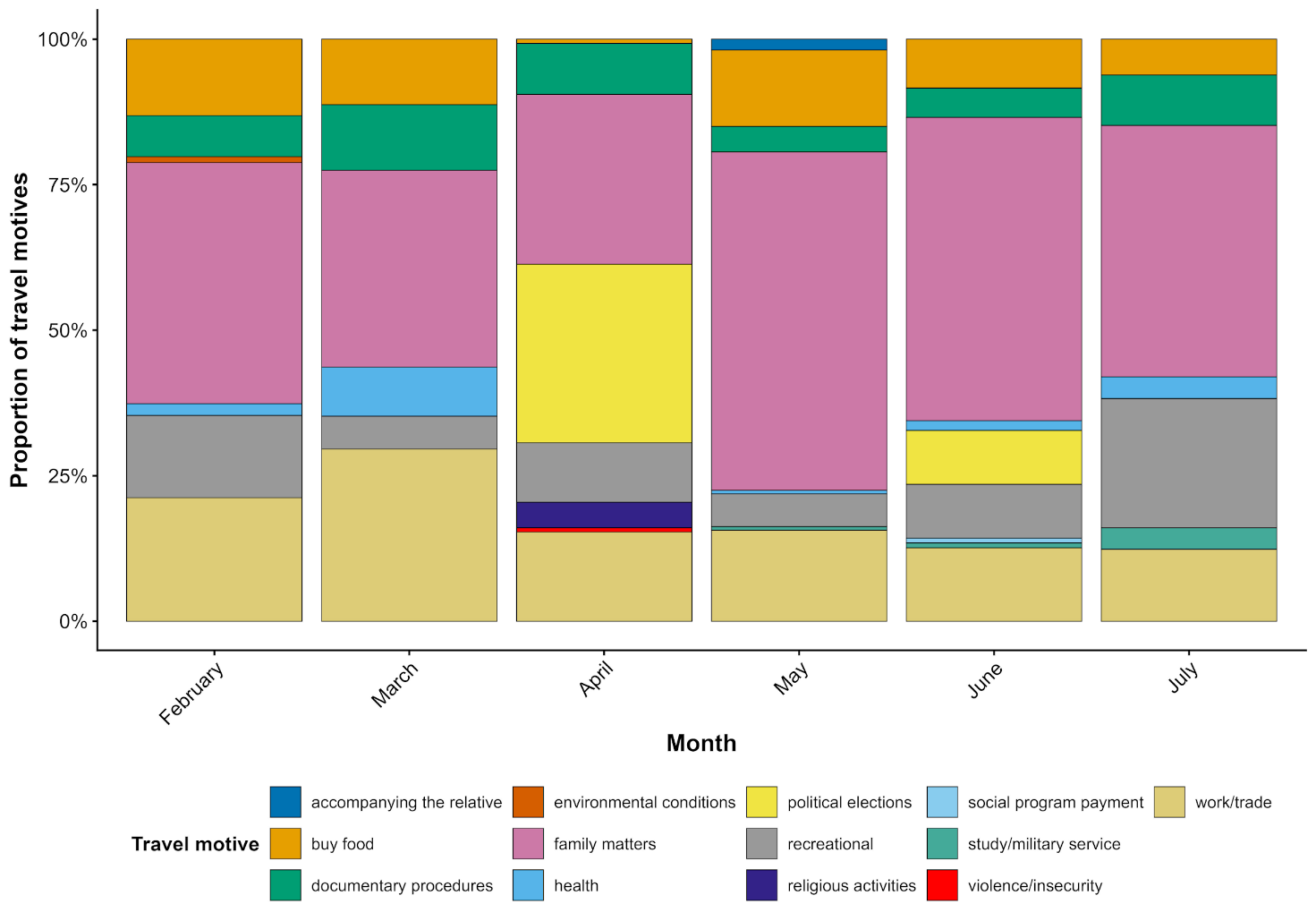

**Supplementary Figure 1.** Proportion of reasons for internal mobility among individuals from the communities of Libertad (A) and Urcomiraño (B).

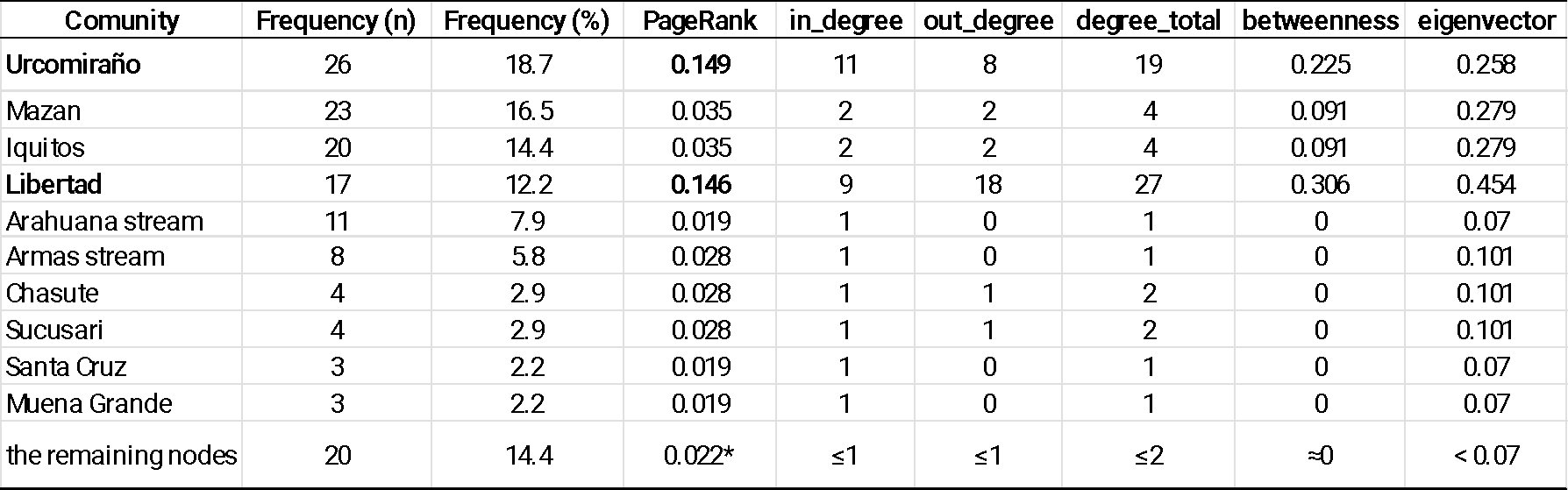

* Approximate average PageRank of the set of peripheral nodes

**Supplementary Table 2.** Travel frequency vs. Centrality (PageRank) and structural metrics
